# Household clusters of SARS-CoV-2 Omicron subvariants contemporaneously sequenced from dogs and their owners

**DOI:** 10.1101/2024.12.02.24318339

**Authors:** Francisco C. Ferreira, Lisa D. Auckland, Rachel E. Busselman, Edward Davila, Wendy Tang, Ailam Lim, Nathan Sarbo, Hayley D. Yaglom, Heather Centner, Heather Mead, Ying Tao, Juan Castro, Yan Li, Jing Zhang, Haibin Wang, Lakshmi Malapati, Peter Cook, Adam Retchless, Suxiang Tong, Italo B. Zecca, Ria R. Ghai, Casey Barton Behravesh, Rebecca S. B. Fischer, Gabriel L. Hamer, Sarah A. Hamer

## Abstract

Monitoring the zoonotic potential of emerging SARS-CoV-2 variants in animals is a critical tool to protect public health. We conducted a longitudinal study in 47 households reporting people with COVID-19 in Texas in January-July 2022, during the first Omicron wave. We evaluated 105 people and 100 of their companion animals by RT-qPCR for SARS-CoV-2 at three sequential sampling events 1-2 weeks apart, starting 0-5 days after the first reported diagnosis of COVID-19 in the house. Of 47 households that reported people with COVID-19, SARS-CoV-2 was detected in 43, with 68% of people testing positive by RT-qPCR; 95.5% of people had antibodies to SARS-CoV-2. Dogs were the only animal species positive by RT-qPCR (5.4%; 3/55). Viral copies were consistently lower in dogs than household members, and no infectious virus was recovered in cell culture. Whole genome sequencing revealed household clusters of Omicron subvariants BA.1.1, BA.2.3.4 and BA.5.1.1 in people, dogs and a food bowl, confirming human-to-dog transmission within households, with no evidence of onward transmission from the infected dogs. Eleven dogs (n = 55) and two cats (n = 26) had neutralizing antibodies against SARS-CoV-2. Infection was not associated with clinical signs in pets; only two animals that tested negative for SARS-CoV-2 were reported to be sick. Nearly one-third (30.2%) of households with active COVID-19 had pets exposed to SARS-CoV-2, similar to our pre-Omicron studies, yet incidence of infection in cats was lower compared to pre-Omicron. These differences suggest that the zoonotic transmission dynamics in households may differ based on variant.

**Significance statement:** Monitoring companion animals offer insights into the zoonotic potential of SARS-CoV-2 ahead of its introduction into other animal populations where viral spread may go unchecked. At the peak of the first Omicron wave, we assessed SARS-CoV-2 transmission dynamics in households longitudinally testing people and their pets in Texas. Omicron infections in cats were significantly lower when compared to pre-Omicron variants. Whole genome sequencing revealed three household clusters of human-to-dog transmission, each with a different Omicron subvariant, yet we did not find evidence of onward transmission to other animals or humans from infected dogs. Sustained animal surveillance in at-risk animals and people using the One Health approach are critical given the ongoing potential for viral evolution that can impact public health.

## Introduction

Severe acute respiratory syndrome virus 2 (SARS-CoV-2), the agent of coronavirus disease 2019 (COVID-19), is a zoonotic virus of worldwide importance to public health and to the health of at-risk mammalian species. Spillback infections from people to animals are widespread given the wide range of susceptible mammal species (1). Confirmation of the susceptibility of companion animals, especially cats and dogs, to SARS-CoV-2 via experimental challenges (2, 3) and the detection of natural infections at early stages of the COVID-19 pandemic across multiple continents (4–6) raised questions about the role of pets in transmission cycles of the virus. However, few human infections resulting from contact with infected pets have been reported (7, 8).

The highly transmissible Omicron variant emerged globally in November 2021, and confirmed predictions that virus variants and subvariants may differ in their host-range (9, 10). Because dogs and cats remained susceptible to Omicron under laboratory (11, 12) and in natural conditions (13, 14), it is essential to survey pets for their involvement in SARS-CoV-2 transmission cycles as new variants continue to emerge. This is critical because dogs experimentally infected with Omicron can sustain onward transmission to naive conspecifics (12).

Pets living in households with active COVID-19 cases are more likely to have been exposed to SARS-CoV-2 compared to pets with no or unknown evidence of exposure to infected people (15–17). While cross-sectional studies are useful for determining pet infection prevalence, longitudinal studies of pets and household members are needed to understand transmission dynamics within households. Therefore, we conducted a longitudinal investigation enrolling people and pets living in households with active COVID-19 cases during the emergence of Omicron in Texas to understand inter-species transmission patterns under natural conditions.

## Results

### General results

In total, we sampled 105 people and 100 animals from 47 households where at least one person self-reported COVID-19, with 1-7 people per house and 1-12 pets per house (Table 1). Thirty-five households had dogs, 19 had cats and nine had both dogs and cats. An average of 1.63 dogs (n = 57; 0.88 standard deviation) and 1.38 cats (n = 29; 0.59 SD) were tested per household. The mean age for dogs and cats was 6.7 (4.58 SD) and 6.4 (3.99 SD) years, respectively. The cohort also included five goats, three horses, two pigs, a donkey, a rabbit, a gecko and a tortoise.

**Table 1:**
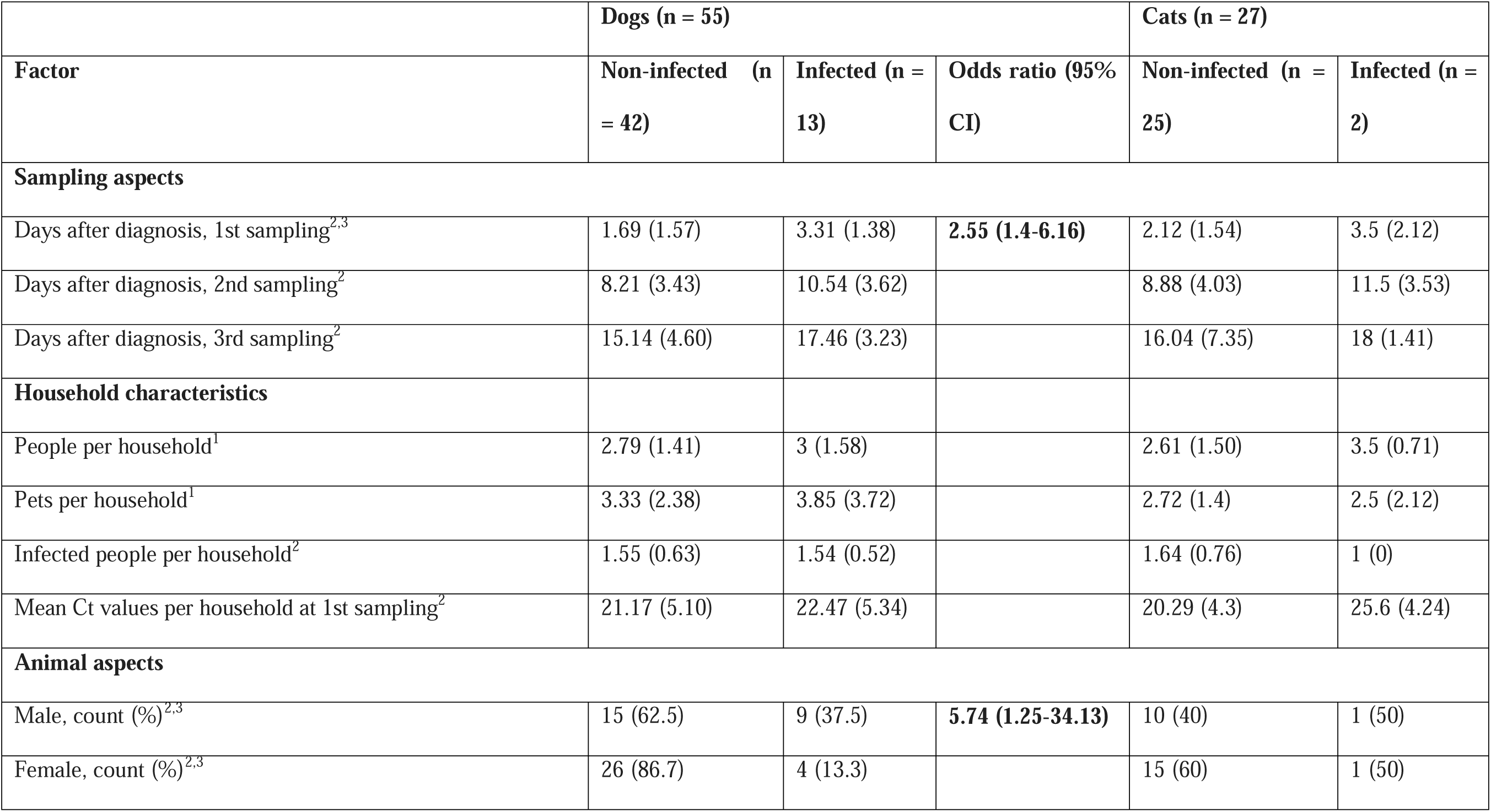
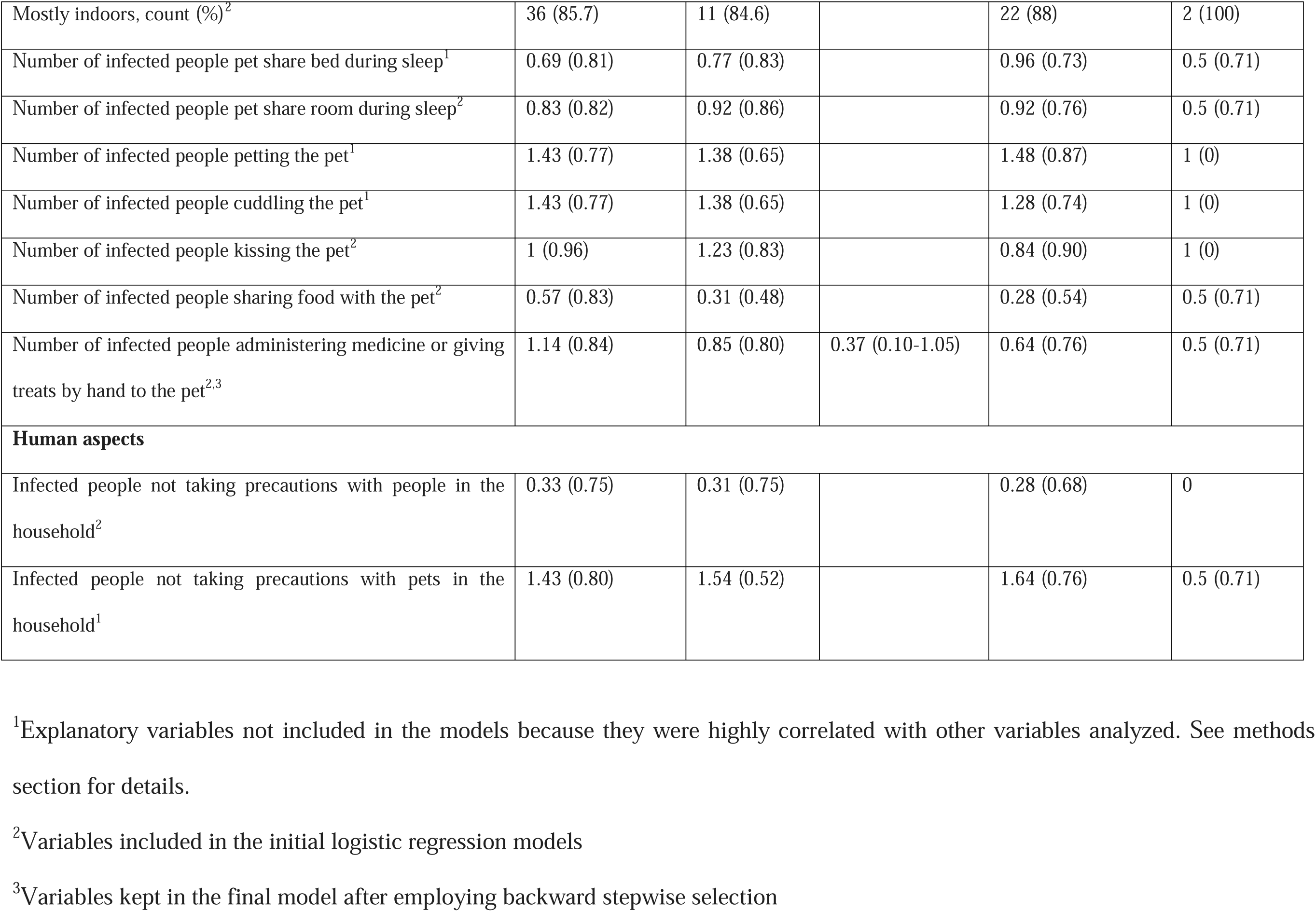
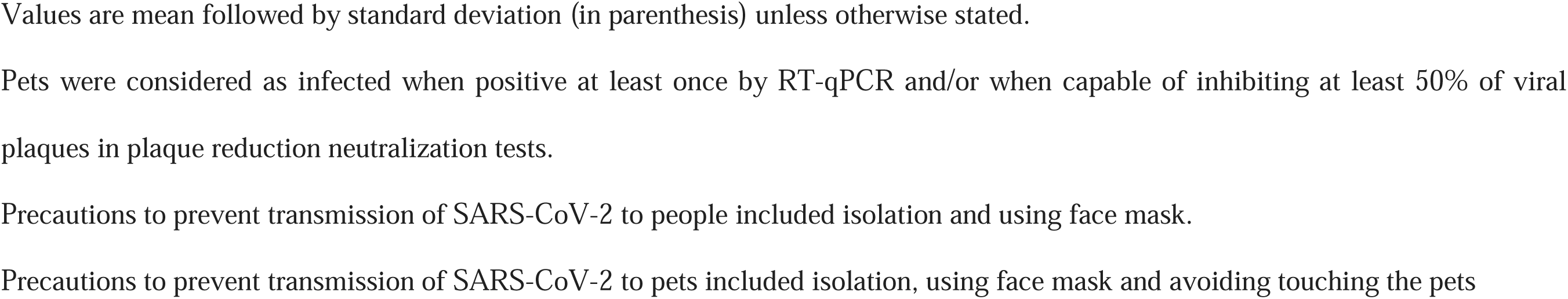
General statistics for 43 households in Texas with active SARS-CoV-2 infections in humans owning pets. Logistic regression models were used to determine factors associated with risk of infection in dogs only due to the small numbers of cats sampled and determined as infected with or exposed to SARS-CoV-2. Odds ratio and 95% confidence intervals were included for the factors kept in the final model; significant values are in bold.

We collected swabs at three consecutive sampling events 0-33 days after the self-reported date of first COVID-19 diagnosis within each household (referred to as “days after diagnosis”; Table 1). The average number of days after diagnosis was 2.22 (1.6 SD) for the first sampling, 8.9 (3.7 SD) for the second, and 15.7 (5 SD) for the third.

### Detection of SARS-CoV-2

We confirmed at least one positive result among people with SARS-CoV-2 via RT-qPCR in 43 of the 47 households; households with no SARS-CoV-2 detection in humans were excluded from analyses below. Positivity rate by RT-qPCR in humans (samples positive by both N1 and N2 tests) was 63.9% (n = 97), 52.1% (n = 96), 22.6% (n = 93) at the first, second and third sampling event, respectively (Supplementary Figure S1). Overall, 68.4% (67/98) of the people tested positive at least once. Our generalized linear mixed models (GLMMs) showed that Ct values increased for both N1 and N2 assays with the number of days after diagnosis (*P* < 0.001) between the first and the second sampling event (*P* < 0.001), and between the second and the third sampling event (*P <* 0.001; Supplementary Figure S1).

Of the 55 dogs sampled, three (5.4%) from different households had respiratory swabs positive by RT-qPCR; positive samples were collected at 2, 5, and 9 days after diagnosis of the first person with COVID-19. Rectal swabs from these dogs were negative at all sampling events. All swab samples (respiratory and rectal) from cats (n = 27) and other pet species were negative by RT-qPCR. A total of 3 of 43 households (7%) with people with COVID-19 had RT-qPCR positive pets. Overall, 33 households owned dogs, indicating dogs in 9.1% of these households became positive following potential exposure. The positivity rate by RT-qPCR is lower for cats when compared to our prior, pre-Omicron study ((18) *in preparation*; n = 157; 13.4%; Fisher’s Exact Test, *P* = 0.048), but rates does not differ for dogs (n = 396; 4.8%; Fisher’s Exact Test, *P* = 0.7).

The Ct values from these dogs were consistently higher than values from humans sampled at the same time using the day of the first COVID-19 diagnosis as a reference (GLMM; *P* = 0.008 for the N1 and *P* = 0.007 for the N2 assay; Figure 1). The first two positive dog samples were collected during the first sampling event, while the third dog converted from negative to positive between the first and second sampling. In all cases, the household with a positive dog had a second pet (two with dogs and one with a cat) that remained negative by RT-qPCR in all three sampling events.

**Figure 1.**
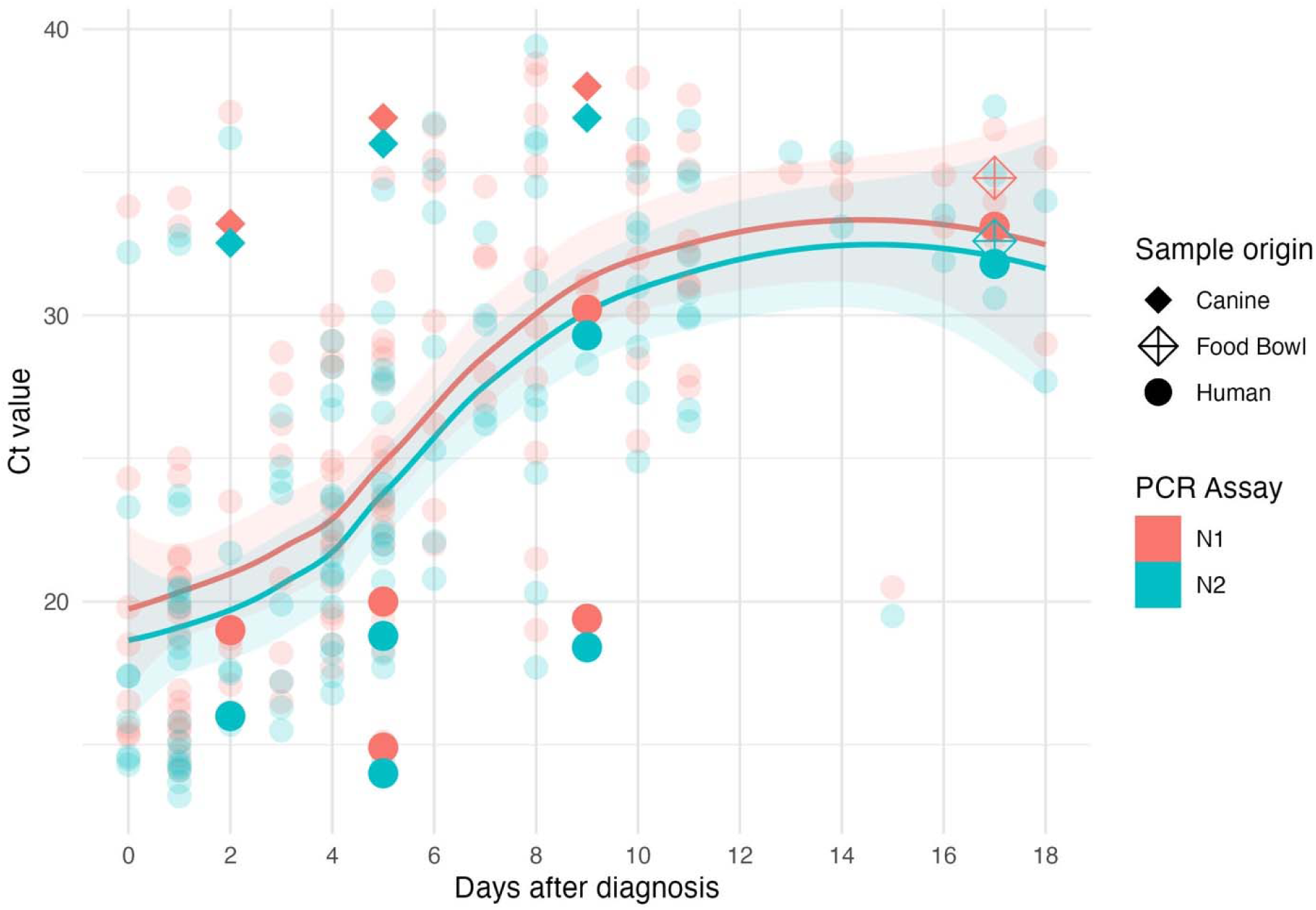
Distribution of SARS-CoV-2 RT-qPCR Ct values from positive people (67 testing positive at least once, with a total of 133 positive samples), dogs (n = 3) and a food bowl in relation to the number of days after diagnosis of first COVID-19 case in the household. Locally estimated scatterplot smoothing (“loess” function) was fitted for data from each RT-qPCR assay for human samples, displaying 95% confidence intervals. Ct values from household members sampled in the same day as positive dogs and the food bowl are in bold colors.

A total of 39 dogs and 21 cats from 32 households had their food and water bowls tested, and a food bowl utilized by a RT-qPCR-positive dog likewise tested positive (2.6%) by RT-qPCR. This household also had a cat that tested negative by RT-qPCR at all three sampling events, and whose food bowl also consistently tested negative.

Attempts to isolate viable virus from RT-qPCR-positive samples collected from dogs and from the food bowl by passages on Vero cells expressing Transmembrane Protease, Serine 2 and Human Angiotensin-Converting Enzyme 2 (Vero E6-TMPRSS2-T2A-ACE2) were unsuccessful. Ct values for these samples ranged from 32.5 to 36.9.

### Whole genome sequencing and phylogeny

Whole-genome sequencing revealed that viruses detected in humans, dogs and a food bowl from each of the three households clustered in monophyletic clades by household (Figure 2).

**Figure 2.**
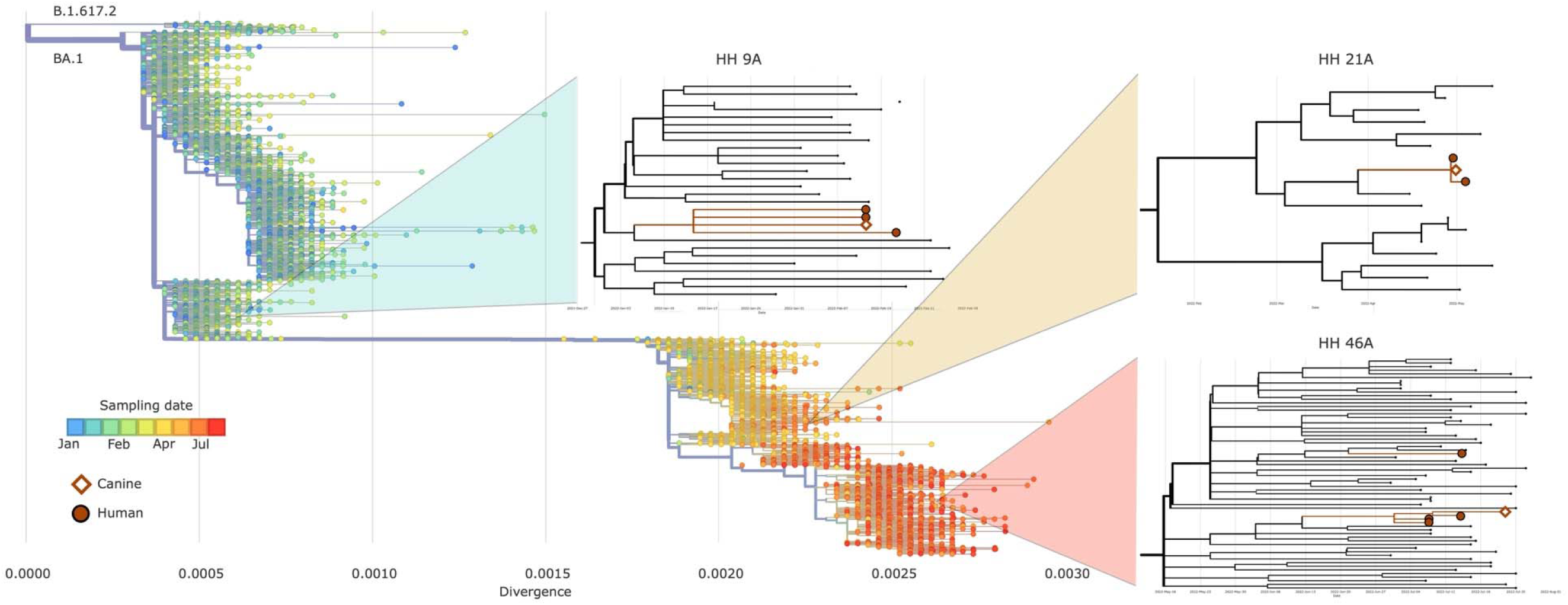
Phylogenetic context of SARS-CoV-2 from dogs in relation to people from the same households. Genome sequences of SARS-CoV-2 from each household with an infected dog were analyzed alongside 15305 sequences from community surveillance in Texas collected within two weeks before or after start of sampling at each household. The main phylogeny at the top left shows sequence divergence with branch labels for the major PANGO lineages and color-coding for the time of specimen collection. Insets show specimens from each household displayed with the most closely related surveillance samples and positioned by collection date. Viral hosts are indicated with markers: circle for human; diamond for canine (including water bowl in HH 46A).

The subvariant BA.1.1 was detected in samples collected from a dog (GISAID acc. num.: EPI_ISL_18065574, GenBank acc. num.: OR398175) and from two people (EPI_ISL_18065564, EPI_ISL_18065569; OR398179, OR398183) living in the same household (HH 9A). Sequencing the virus collected at the second sampling event from one of these two people revealed the same virus sequence (EPI_ISL_18065567; OR398182). Sequences obtained from humans and from the dog had 99.6-99.7% and 95.8% coverage of the SARS-CoV-2 genome, respectively, being identical to each other when excluding gaps in coverage.

In a second household (HH 21A), a dog and a person were infected by the Omicron lineage BA.2.3.4 (EPI_ISL_18065573; OR398173 and EPI_ISL_18065570; OR398184, respectively). We successfully sequenced the same virus genome from this person at the second sampling event (EPI_ISL_18065566; OR398181). Sequence coverage was 99.5-99.6% for human and 98.2% for dog samples, respectively, and were identical to each other.

In the third household (HH 46A), the Omicron lineage BA.5.1.1 was sequenced from two people during the first sampling event (EPI_ISL_18065565, EPI_ISL_18065562; OR398180, OR398177; sequencing coverage = 99.6%); a second viral sequence was obtained from one of the people during the second sampling event (EPI_ISL_18065563; OR398178; sequencing coverage = 99.6%) and from the other person during the third sampling event (EPI_ISL_18065561; OR398174; sequencing coverage = 90.1%). The virus sequences obtained during the first two sampling events were identical to the one obtained from a dog food bowl that was positive in the third sampling event (EPI_ISL_18065572; OR398176; sequencing coverage = 93.1%). The virus sequenced from one person sampled in the third event, 14 days after the first sampling, had six single nucleotide polymorphisms (SNP) and a six-nucleotide deletion compared to the other sequences obtained from this household. One of these mutations (C27532A), which was not present in the first sample sequenced from this person, placed this sequence into a separate clade within BA.5.1.1 from Texas, suggesting an independent infection event. The virus detected from the dog was not successfully sequenced. Shotgun metagenomic sequencing was conducted to verify the species origin of samples from the positive food bowl from this household, which showed a mixture of mitochondrial DNA matching human and canine, but also chicken, cow, and pig, likely reflecting dog food components.

### Serology

We used an ELISA assay targeting IgG against SARS-CoV-2 spike protein (Inbios, Seattle, USA) in eluates from dried blood spots (DBS) collected from people in the first sampling event. This serological test detects IgG in response to either natural exposure to SARS-CoV-2 or vaccination. Overall, 95.3% of the people (82/86) were seropositive. Five people with serology data (5.8%) reported never having been vaccinated against COVID-19, three of which were seropositive and tested positive by RT-qPCR, at days 3, 5, and 5 after the initial COVID-19 diagnosis in their households. Only one person was vaccinated yet tested negative by serology.

Eleven out of 55 dogs (20%) had antibody titers capable of neutralizing the formation of at least 50% of viral plaques (PRNT_50_-positive), two of which (18.2%) neutralized 90% or more virus plaques (PRNT_90_) at 1:10 and at 1:20 dilutions during all three sampling events. Two cats (7.7%; n = 26) were PRNT_50_-positive (Figure 3a). Five goats, three horses, and one donkey tested negative by PRNT. Two households had two pets each that were seropositive, while seven other multi-pet households had only a single seropositive animal (Figure 3a). SARS-CoV-2 active infection and/or past exposure in pets, as demonstrated by positivity by RT-qPCR and by PRNT, respectively, was not statistically different between dogs (23.6%) and cats (7.4%; Fisher’s Exact Test, *P* = 0.13). Seroprevalence is lower for cats when compared to our pre-Omicron study (n = 146; 35.7%; Fisher’s exact test, *P* = 0.005), but rates are similar for dogs (n = 382; 24.9%; χ^2^ with Yates correction = 0.4, *P* = 0.54; (18)).

**Figure 3.**
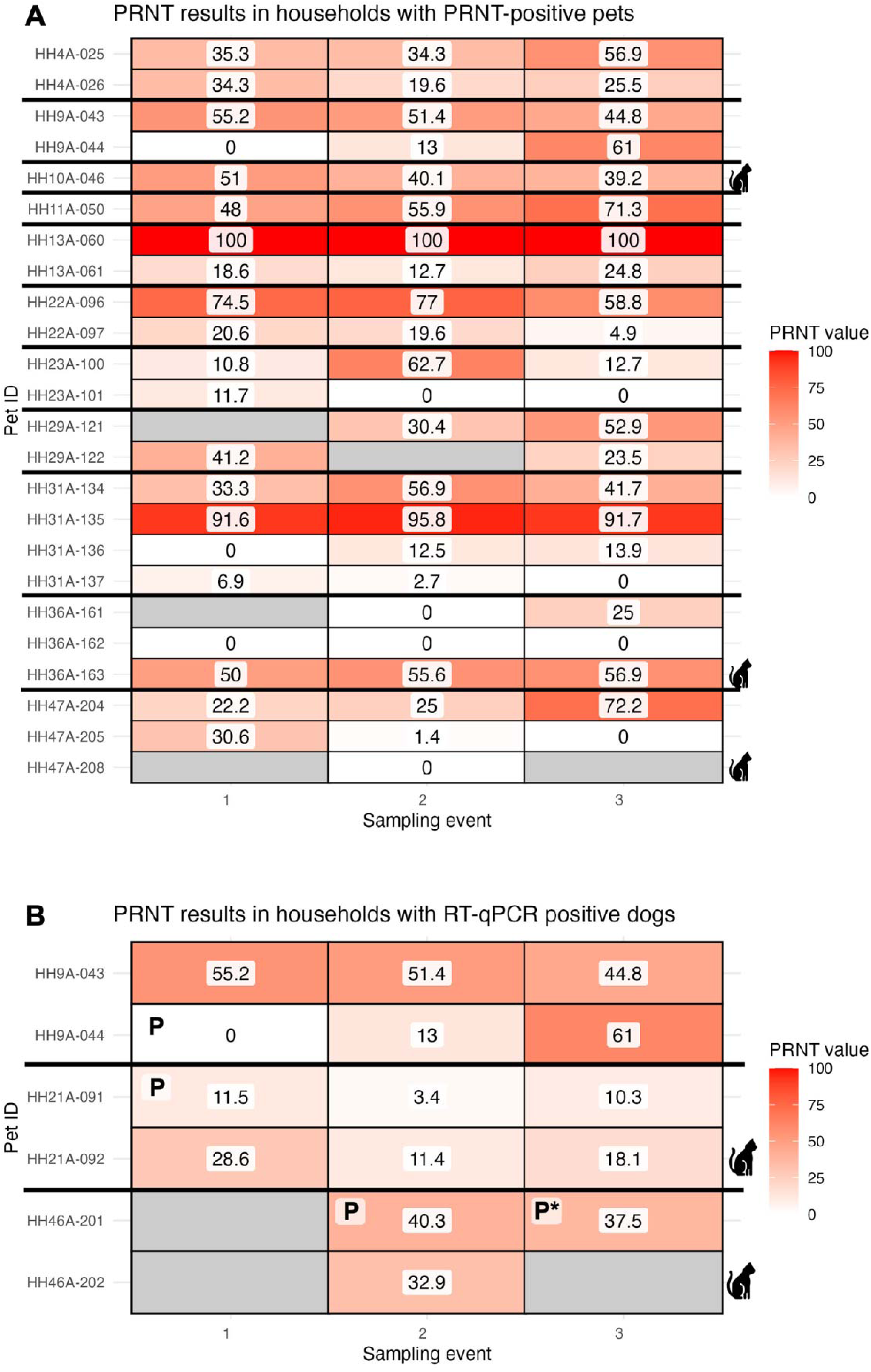
Plaque reduction neutralization assay (PRNT) results over time for dogs and cats sampled in households with a human COVID-19 case. Numbers inside boxes represent the proportion of viral plaques neutralized per pet per sampling event. **A.** All pets from households with seropositive pets at one or more time points are displayed. **B.** PRNT_50_ for pets in households with RT-qPCR-positive dogs, which are identified with a “P”. The food bowl from dog HH46A-201 was RT-qPCR-positive at the third sampling point (“P*). Households are separated by thick horizontal lines; tiles in grey indicate missing data. Cats are indicated by icons; all other individuals are dogs.

The dynamics for RT-qPCR and PRNT_50_ tests for pets positive by RT-qPCR and for other pets from the same household are shown in Figure 3b. Only one RT-qPCR-positive dog had neutralizing antibodies (PRNT_50_ positive) against SARS-CoV-2, which were first detected at the third sampling event. The other dog from this same household was PRNT_50_-positive at the first and second sampling events. None of the other RT-qPCR-positive dogs or pets with which they were co-housed were PRNT_50_-positive.

We examined viral plaque neutralization capacity over time among samples that did not meet the PRNT_50_ threshold. At the first, second, and third sampling events, mean PRNT values were 21.9% (SD = 21.8), 24.2% (SD = 24.2), and 27.1% (SD = 23.6) for dogs, and 20.4% (SD = 16.3), 21.2% (SD = 14.6) and 16.2% (SD = 14.4) for cats, respectively, with no difference over time.

Combining both RT-qPCR and serostatus data, infection rates are lower for cats when compared to our pre-Omicron study (n = 157; 35.7%; Fisher’s Exact Test, *P* = 0.002) but are similar for dogs (n = 396; 27.3%; χ^2^ with Yates correction = 0.6, *P* = 0.68). Overall, 13 out of 43 (30.2%) households with active COVID-19 cases had pets infected/exposed to SARS-CoV-2, similar rates to pre-Omicron (n = 281; 39.1%; χ^2^ with Yates correction = 1, *P* = 0.31).

### Survey and odds of infection in pets

The only two pets reported by household members to have clinical signs of disease (coughing and lethargy) were dogs that tested negative by both diagnostic methods at all three time points. Logistic regression models indicated that the odds of detecting infected or exposed dogs were correlated with the number of days after human diagnosis in the first sampling event (Table 1; *P* = 0.01). Specifically, for each additional day after human diagnosis, the odds of being positive increased by a factor of 2.55 (95% CI: 1.40 - 6.16). No correlation was observed for the second and third sampling events. Male dogs had 5.74 higher odds of being positive compared to females (95% CI: 1.25 - 34.14; *P* = 0.03). Although the number of infected people administering medicine or providing treats to dogs showed a trend towards reducing the odds of positivity (odds ratio = 0.37, 95% CI: 0.10 - 1.05), this effect was not statistically significant (P = 0.09). None of the other explanatory factors included in the initial models were retained in the final model (Table 1).

## Discussion

We conducted a longitudinal study using a One Health approach by examining humans and their pets for SARS-CoV-2 infection. Our study from January 2022 through July 2022, took place during the peak of the BA.1 Omicron wave to the BA.2/BA.5 wave. We detected SARS-CoV-2 RNA and neutralizing antibodies at low rates in pets, particularly cats, despite multiple sampling starting shortly after human COVID-19 diagnosis (average of 2.2 days). We show transmission dynamics within houses, with the conversion of a dog from RT-qPCR-negative to positive between consecutive sampling events six days apart, emphasizing the importance of longitudinal sampling. By obtaining full virus genomes from people, pets and from a food bowl utilized by a RT-qPCR-positive dog, we confirmed horizontal transmission from humans to their pets within households.

Within the three households with RT-qPCR-positive dogs, all co-housed pets remained negative despite sequential sampling. This is especially remarkable given lower detection rates in cats in this study compared to pre-Omicron (18). Our results are consistent with a prior study reporting a marked reduction in seropositivity in pets after the emergence of the Omicron variant (19).

The Ct values in dogs were consistently above the upper limit of the confidence interval of Ct values from human samples collected at the same reference time, suggesting lower viral loads in dogs. Ct values may or may not be negatively correlated with the likelihood of detecting infectious virus in biological samples (reviewed by (20)), but our results provide epidemiological data showing that the low infectability of dogs during the high exposure time period, and lack of onward transmission to other pets may be associated with high Ct values during active infections. Failure to isolate virus from RT-qPCR-positive swab samples from pets further confirm that they were not shedding or shedding extremely low levels of infectious virus at the time of sampling. These samples were collected between two and nine days after diagnosis of the human cases. We, therefore, suggest that Omicron variants are not efficiently transmitted from humans to pets or between pets under natural conditions. Factors such as increased levels of immunity due to vaccines and prior infection could have contributed to reduced human-to-pets transmission.

Of note, 49% of dogs and 48% of cats were living with between two and four people with active COVID-19 that were not taking precautions to prevent SARS-CoV-2 transmission to their pets in most cases, revealing high chances of natural exposure to the virus. However, none of the factors related to dog-human interactions were associated with increased odds of infection. For instance, sharing food with pets has been associated with increased human-to-animal transmission of SARS-CoV-2 (21), but we did not detect such effect. However, the small number of households and pets tested may have limited the power of our analysis.

Genomic epidemiology has inferred only two and four cases of transmission of SARS-CoV-2 from dogs and cats to humans, respectively (22). However, the inferred number of transmissions from humans to pets was at least 13 times higher than those values. Our results provide evidence that pets may not transmit the Omicron variants efficiently intra- and inter- species under natural conditions, which would be one of the factors explaining the low likelihood of pet-to-human transmission (22).

Whole genome sequencing confirmed that dogs and people from the same household were infected by the same virus in two cases, with no variation in the viral sequences recovered from the animals vs. the people. Additionally, we saw that the same virus infecting people was present in the food bowl from a household with a positive dog. One specimen from this household contained a viral sequence with six SNPs and a deletion relative to the other viral sequences from the household, including one sequence obtained from the same individual two weeks earlier. This number of mutations is at the extreme end of what is typically observed among viruses collected from the same household (23) or in persistent infections (24), suggesting that this may be a separate introduction of SARS-CoV-2 into the household. This inference is supported by one of these mutations being shared with sequences obtained from community surveillance activities in Texas in the weeks prior. Reinfections within a period of a few weeks are rare, but have been reported (24).

Our detection of a RT-qPCR-positive dog food bowl may reflect use by a positive dog and/or contamination by a positive household member. People experimentally infected with the wild-type SARS-CoV-2 contaminated household items with the virus (25). Moreover, contaminated surfaces may be correlated with increased within-household transmission risk (26). Both people from the household with the positive food bowl were positive, with average Ct values of 16.9 at the first sampling event. However, the food bowl tested positive only at the third sampling, when only one person was still positive (average Ct = 32.5), while the dog was positive at the second sampling event only (average Ct = 37.5). This suggests that viral RNA detected in the food bowl was derived from the positive dog.

Experimentally infected cats shed lower virus loads of an Omicron variant when compared to pre-Omicron variants (11). Additionally, a recent study did not isolate infectious virus from cats infected with a low dose of Omicron, despite recovering infectious viruses from cats infected with the ancestral, gamma and delta strains (27). This helps to explain the lack of SARS-CoV-2 detection in cats in our study, and the lower detection rates here when compared to a broad study conducted pre-Omicron also in households with active COVID-19 cases in Texas (18).

In experimental challenges, cats seroconvert at 7 days post challenge with pre-Omicron strains, and dogs seroconvert 7-14 days (2, 11), while Omicron-infected cats display a delayed seroconversion between 7 and 14 days post infection (11) or do not seroconvert when exposed to low viral doses (27). The repeated nature of our study allowed the detection of late seroconversion in seven out of 11 seropositive dogs. We also observed that most seropositive pets had low antibody titers (PRNT_50_-positive at 1:10 serum dilution only). These low titers could explain the fact that 30.1% of these pets were seronegative in a subsequent sampling event, suggesting waning neutralizing antibodies within one to two weeks. All but one of these pets were negative by RT-qPCR, indicating that the low titers may alternatively reflect exposures to SARS-CoV-2 earlier in the pandemic. Lack of seroconversion in two of the three infected dogs, including one that was resampled five and eleven days after the positive RT-qPCR diagnosis, and the other resampled eight days after RT-qPCR positivity, suggest late seroconversion or undetectable titers of neutralizing antibodies.

### Conclusions

Multiple Omicron variants were detected in household clusters among dogs and household members, yet infection rates in cats were lower than in pre-Omicron studies. The high Ct values in RT-qPCR assays targeting SARS-CoV-2 and lack of infectious virus in samples from dogs, associated with a lack of evidence of onward transmission between pets, indicate that dogs and cats were unlikely to act as amplifying hosts for early Omicron variants. As host-breadth and virus fitness change with the evolution of new variants, continued surveillance using One Health approaches may be critical as new waves driven by a diversity of viruses are expected for years to come. Involving companion animals in SARS-CoV-2 surveillance efforts may inform if and how the virus can affect other animals populations, and may serve as a proxy when humans cannot be sampled directly.

## Methods

### Recruiting and sampling

Between January-July 22, 2022, we recruited people and pets living in the same household as a person with a SARS-CoV-2 infection via COVID-19 portal of Texas A&M University (TAMU). Participating persons responded to a short questionnaire by phone to tally the number of humans and pets living in the house; human COVID-19 vaccination history, date of positive human test result, pet species and signalment (breed, age, sex), and any pet clinical signs of disease. The TAMU Institutional Animal Care and Use Committee (2018-0460 CA) and Clinical Research Review Committee approved animal sampling. The Institutional Review Board issued a public health surveillance exemption.

We visited each house three times over an approximate 2-week period for sample collection. We collected nasal swabs from people during all three sampling visits. From the pets, we collected nasal and oral swabs that were combined into a single vial containing 3 mL of viral transport media (VTM; made following CDC SOP#: DSR-052-02), while rectal swabs were stored in a separate vial with 3 mL of VTM. Additionally, in a subset of households, we collected swab samples from food and water bowls utilized by the pets. Swabs in VTM were stored in a cooler with ice packs until arrival in the laboratory, where samples were stored in a −80 °C freezer.

From humans, we collected blood onto Whatman® protein saver cards (Sigma-Aldrich, St. Louis, MO, USA) via finger prick at the first visit only. These samples were air-dried and stored at room temperature. From pets, blood was collected from either jugular, cephalic or saphenous veins into clot activator tubes and kept in a cooler until centrifugation with serum aliquots stored at −80°C.

### Molecular testing

VTM aliquots from humans, pets, food and water bowls were shipped to the Wisconsin Veterinary Diagnostic Laboratory for RT-qPCR targeting nucleocapsid gene region 1 (N1) and nucleocapsid gene region 2 (N2) (28, 29) of SARS-CoV-2. Cycle threshold (Ct) values were reported as the average of the values for regions N1 and N2.

### Virus isolation

VTM aliquots of three pets and the food bowl that tested positive by RT-qPCR were transferred to a BSL-3 laboratory in Texas A&M University. For virus isolation, 100 µL of VTM with 900 μL of 1× Dulbecco’s Modified Eagle Medium (DMEM) via syringe filtration using an 0.2 micron pore size onto Vero E6-TMPRSS2-T2A-ACE2 (BEI Resources, NR-54970) cells expressing both endogenous cercopithecine ACE2 and TMPRSS2 as well as transgenic human ACE2 and TMPRSS2. Plates were incubated for 72 h and presence of cytopathic effects was evaluated using a brightfield microscope following published protocols (30).

### SARS-CoV-2 whole genome sequencing, metagenomics, and phylogenetic analysis

Aliquots of VTM from infected pets, food bowl, and the humans living with infected pets were sent to the CDC for SARS-CoV-2 whole genome sequencing according to established protocols (31). Phylogenetic tree were inferred using RAxML in the NextStrain pipeline (v7.1.0; (32)) with all 15305 SARS-CoV-2 genomes from the GISAID database that were detected in Texas two weeks before or after the start of sampling at each household. PANGO lineages were assigned using ‘pangolin’ (software v4.3.1; data v1.29; (33)). Mammalian mitochondrial DNA was identified by untargeted metagenomic sequencing as described previously (34) followed by mapping reads to a database of mtDNA sequences representing clusters with 93% sequence identity (35).

### Serologic testing

At the TGen North laboratory, human dried blood spots were eluted to 1:100 in dilution buffer. Anti-SARS-CoV-2 antibodies were detected using a qualitative SARS-CoV-2 IgG ELISA assay (InBios, Seattle, WA, USA). All assays, including controls, were used as per the manufacturer’s recommendations.

Pet serum samples were tested by plaque neutralization tests (PRNT) at Texas A&M University Global Health Research Complex to quantify neutralizing antibodies against SARS-CoV-2 in BSL-3 following the protocol described by Roundy et al. (36). Briefly, we used Vero CCL-81 cell cultures in 6-well plates and SARS-CoV-2 isolate USAIL1/2020, NR 52381 (BEI Resources, Manassas, VA, USA) for an initial screening using serum samples at a dilution of 1:10 to test their ability to reduce virus plaques by at least 50% when compared to the virus control, a well-established method (37) with sensitivity of 97% (38) which reduced the chances of assigning pets as false-negative in our study. The subset of positive serum samples with antibody titers able to reduce more than 90% of virus plaques were further tested at 2-fold serial dilutions to determine 90% endpoint titers.

### Statistical analysis

We used either Chi-squared test with Yates’ correction or Fisher’s Exact test to compare positivity rates by RT-qPCR and PRNT_50_ between this study and a previous one conducted pre-Omicron in the same region in Texas (18).

We employed generalized linear mixed models (GLMMs) (using the *lme4* package (39)) to analyze changes in Ct values from positive samples using species (humans or dogs), days after diagnosis and sampling event as fixed effects, and host ID as random. We used separate models for N1 and N2 genes, which produced similar results. Given the non-normal distribution of Ct values, we applied a Gamma distribution with a log link function, which produced a better-fit model. To assess changes in PRNT values over time, we employed a GLMM adding species as explanatory variable, also using animal ID as a random effect. We used a Tweedie distribution with a log link function because PRNT values presented a non-normal distribution and had values equal to zero. We performed backward stepwise selection using Akaike Information Criterion (AIC) to identify significant predictors.

We built a logistic regression model to determine factors associated with the risk of dogs becoming infected with SARS-CoV-2. We detected high multicollinearity (correlation values above or below 0.85 and −0.85, respectively) among some explanatory variables. For example, the number of infected people petting, cuddling and not taking precautions to not transmit SARS-CoV-2 were highly correlated with the number of infected people per household. Therefore, only the latter variable was kept. Similarly, the number of infected people sharing the bed and room with pets were also correlated and only the latter was included. The initial model included number of days after diagnosis, pet sex, number of infected people in household, mean SARS-CoV-2 Ct value of people in household, number of infected people interacting with dog (sleeping in same room, kissing, sharing food, giving medicine and treats by hand) and whether the pet stayed >75% indoors. We performed backward stepwise selection using AIC to identify significant predictors in these models. The coefficients from the final model were used to calculate odds ratios (OR) and their 95% confidence intervals (CI). We did not build models for cats because only two individuals tested positive. We performed all analyses using R 4.2.2 (40).

## Data Availability

All data produced in the present study are available upon reasonable request to the authors.

## Acknowledgments

Funding provided by the Centers for Disease Control and Prevention. We thank Dr. Benjamin Neuman and Tahmina Pervin, Texas A&M University, for virus isolation efforts. We thank Dan Christensen from the Wisconsin Veterinary Diagnostic Laboratory for diagnostic support.

## Disclaimer

The findings and conclusions in this report are those of the author(s) and do not necessarily represent the official position of the Centers for Disease Control and Prevention.

---

**Supplementary Figure S1.**
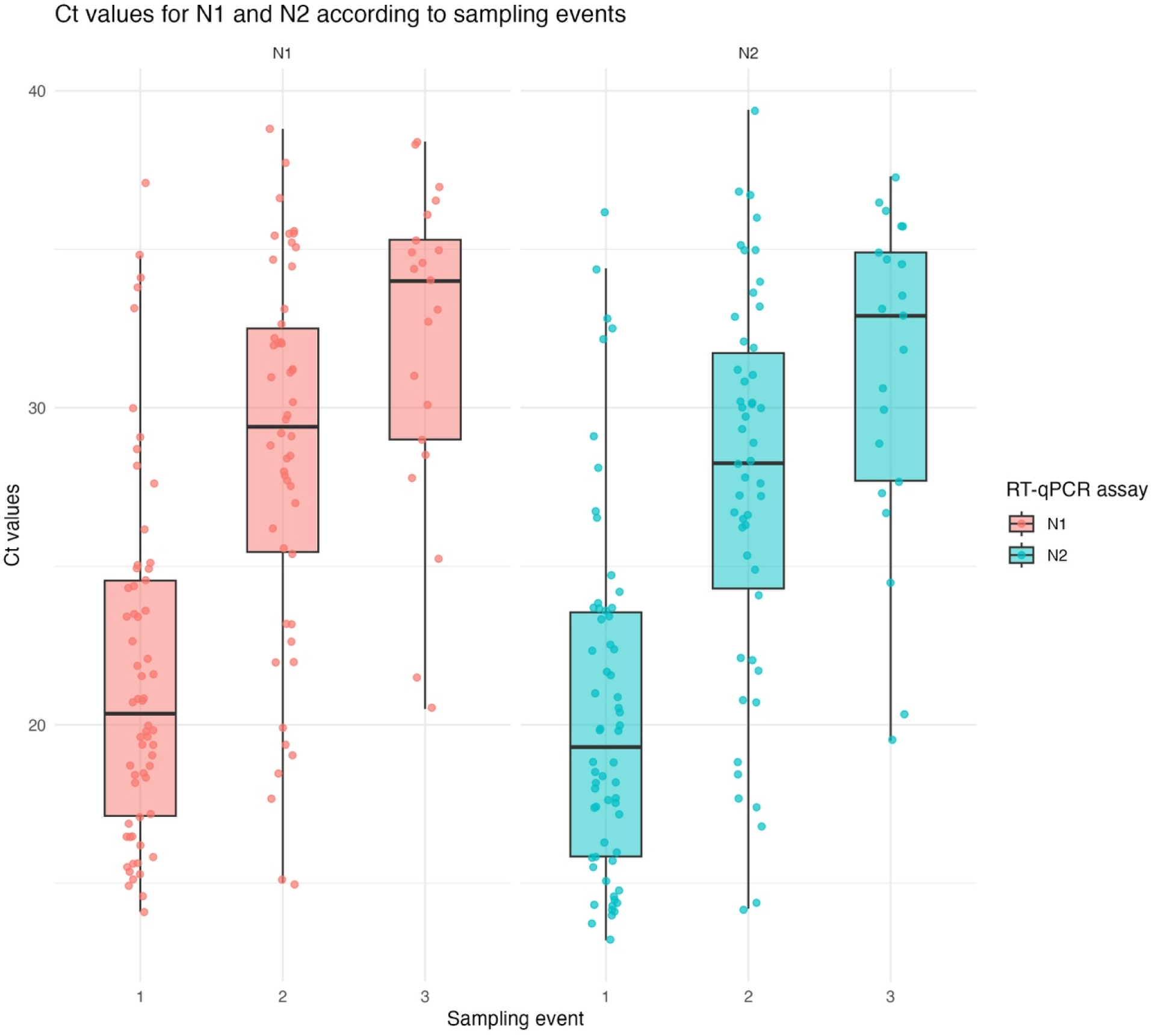
Distribution of Ct values for the two RT-qPCR assays (N1 and N2) from human samples according to sampling event. Median, interquartile range (IQR), and whiskers representing 1.5 times the IQR are displayed. Number of positive samples for the first, second and third sampling points were 62, 50 and 21, respectively. All samplings for event 1 occurred within 0-5 days after the first COVID-19 diagnosis, within 4-18 days for event 2, and within 8-33 days for event 3.

**Supplementary Figure S2.**
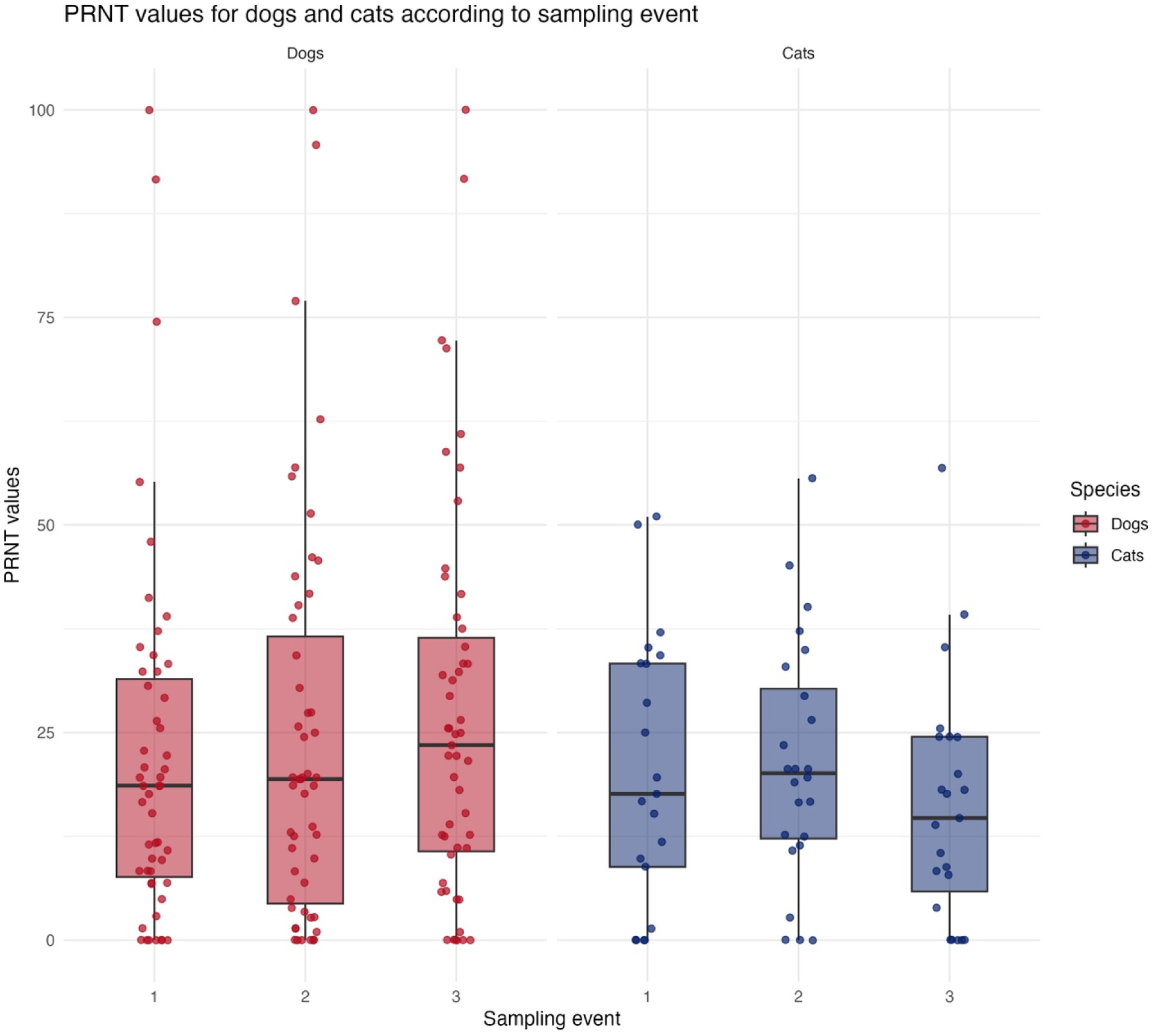
Distribution of PRNT values for samples collected from dogs and cats according to sampling event. Median, interquartile range (IQR), and whiskers representing 1.5 times the IQR are displayed . A total of 55 dogs were tested at least once, with 51, 51 and 51 samples evaluated in the first, second and third sampling event, respectively. A total of 26 cats were tested, with 21, 24 and 23 samples evaluated in the first, second and third sampling event, respectively.

## References

1. WOAH (2023) World Organisation for Animal Health - SARS-COV-2 in animals - situation report 22.

2. A. M. Bosco-Lauth et al., Experimental infection of domestic dogs and cats with SARS-CoV-2: Pathogenesis, transmission, and response to reexposure in cats. Proc Natl Acad Sci U S A 117, 26382–26388 (2020).

3. J. Shi et al., Susceptibility of ferrets, cats, dogs, and other domesticated animals to SARS–coronavirus 2. Science 368, 1016–1020 (2020).

4. Q. Zhang et al., A serological survey of SARS-CoV-2 in cat in Wuhan. Emerging Microbes & Infections 9, 2013–2019 (2020).

5. A. Newman et al., First Reported Cases of SARS-CoV-2 Infection in Companion Animals - New York, March-April 2020. MMWR Morb Mortal Wkly Rep 69, 710–713 (2020).

6. E. I. Patterson et al., Evidence of exposure to SARS-CoV-2 in cats and dogs from households in Italy. Nature Communications 11, 6231 (2020).

7. T. Sila et al., Suspected Cat-to-Human Transmission of SARS-CoV-2, Thailand, July-September 2021. Emerg Infect Dis 28, 1485–1488 (2022).

8. H. L. Yen et al., Transmission of SARS-CoV-2 delta variant (AY.127) from pet hamsters to humans, leading to onward human-to-human transmission: a case study. Lancet 399, 1070–1078 (2022).

9. Z. Niu et al., N501Y mutation imparts cross-species transmission of SARS-CoV-2 to mice by enhancing receptor binding. Signal Transduct Target Ther 6, 284 (2021).

10. G. T. Barut et al., The spike gene is a major determinant for the SARS-CoV-2 Omicron-BA.1 phenotype. Nat Commun 13, 5929 (2022).

11. M. Martins et al., The Omicron Variant BA.1.1 Presents a Lower Pathogenicity than B.1 D614G and Delta Variants in a Feline Model of SARS-CoV-2 Infection. J Virol 96 (2022).

12. K. S. Lyoo et al., Experimental Infection and Transmission of SARS-CoV-2 Delta and Omicron Variants among Beagle Dogs. Emerg Infect Dis 29, 782–785 (2023).

13. C. Piewbang et al., SARS-CoV-2 Transmission from Human to Pet and Suspected Transmission from Pet to Human, Thailand. Journal of Clinical Microbiology 60, e01058–01022 (2022).

14. L. Sánchez-Morales, J. M. Sánchez-Vizcaíno, M. Pérez-Sancho, L. Domínguez, S. Barroso-Arévalo, The Omicron (B. 1.1. 529) SARS-CoV-2 variant of concern also affects companion animals. Frontiers in veterinary science 9, 940710 (2022).

15. L. Fernández-Bastit et al., Severe acute respiratory syndrome coronavirus 2 (SARS-CoV-2) infection and humoral responses against different variants of concern in domestic pet animals and stray cats from North-Eastern Spain. Transbound Emerg Dis (2022).

16. M. Fritz et al., High prevalence of SARS-CoV-2 antibodies in pets from COVID-19+ households. One Health 11 (2020).

17. M. M. Kannekens-Jager et al., SARS-CoV-2 infection in dogs and cats is associated with contact to COVID-19-positive household members. Transbound Emerg Dis 69, 4034–4040 (2022).

18. A. Pauvolid-Corrêa et al. (High prevalence of respiratory shedding of SARS-CoV-2, neutralizing antibodies, and isolations of infectious virus among 580 companion animals from households with a confirmed human COVID-19 case in Texas, United States. In preparation.

19. C. Klein et al., Dogs and Cats Are Less Susceptible to the Omicron Variant of Concern of SARS-CoV-2: A Field Study in Germany, 2021/2022. Transbound Emerg Dis 2023, 1868732 (2023).

20. O. Puhach, B. Meyer, I. Eckerle, SARS-CoV-2 viral load and shedding kinetics. Nature Reviews Microbiology 21, 147–161 (2023).

21. S. Alberto-Orlando et al., SARS-CoV-2 transmission from infected owner to household dogs and cats is associated with food sharing. Int J Infect Dis 122, 295–299 (2022).

22. S. Naderi et al., Zooanthroponotic transmission of SARS-CoV-2 and host-specific viral mutations revealed by genome-wide phylogenetic analysis. eLife 12, e83685 (2023).

23. M. A. P. Donnelly et al., Household Transmission of Severe Acute Respiratory Syndrome Coronavirus 2 (SARS-CoV-2) Alpha Variant-United States, 2021. Clin Infect Dis 75, e122–e132 (2022).

24. M. Ghafari et al., Prevalence of persistent SARS-CoV-2 in a large community surveillance study. Nature 626, 1094–1101 (2024).

25. J. Zhou et al., Viral emissions into the air and environment after SARS-CoV-2 human challenge: a phase 1, open label, first-in-human study. The Lancet Microbe 10.1016/S2666-5247(23)00101-5 (2023).

26. N. Derqui et al., Risk factors and vectors for SARS-CoV-2 household transmission: a prospective, longitudinal cohort study. The Lancet Microbe 4, e397–e408 (2023).

27. E. S. Park et al., The comparison of pathogenicity among SARS-CoV-2 variants in domestic cats. Sci Rep 14, 21815 (2024).

28. G. W. Goryoka et al., One Health Investigation of SARS-CoV-2 Infection and Seropositivity among Pets in Households with Confirmed Human COVID-19 Cases—Utah and Wisconsin, 2020. Viruses 13, 1813 (2021).

29. M. D. Ramuta et al., SARS-CoV-2 and other respiratory pathogens are detected in continuous air samples from congregate settings. Nat Commun 13, 4717 (2022).

30. S. P. Chaki, M. M. Kahl-McDonagh, B. W. Neuman, K. A. Zuelke, Validating the inactivation of viral pathogens with a focus on SARS-CoV-2 to safely transfer samples from high-containment laboratories. Front Cell Infect Microbiol 14, 1292467 (2024).

31. C. R. Paden, et al., Rapid, Sensitive, Full-Genome Sequencing of Severe Acute Respiratory Syndrome Coronavirus 2. Emerg Infect Dis 26, 2401–2405 (2020).

32. J. Hadfield et al., Nextstrain: real-time tracking of pathogen evolution. Bioinformatics 34, 4121–4123 (2018).

33. Á. O’Toole et al., Assignment of epidemiological lineages in an emerging pandemic using the pangolin tool. Virus Evolution 7, veab064 (2021).

34. Y. Li et al., Identification of diverse viruses in upper respiratory samples in dromedary camels from United Arab Emirates. PLoS One 12, e0184718 (2017).

35. A. Crits-Christoph et al., Genetic tracing of market wildlife and viruses at the epicenter of the COVID-19 pandemic. Cell 187, 5468–5482.e5411 (2024).

36. C. M. Roundy et al., High Seroprevalence of SARS-CoV-2 in White-Tailed Deer (Odocoileus virginianus) at One of Three Captive Cervid Facilities in Texas. Microbiology Spectrum 10, e00576–00522 (2022).

37. E. H. Y. Lau et al., Neutralizing antibody titres in SARS-CoV-2 infections. Nature Communications 12 (2021).

38. I. S. Horbach et al., Plaque Reduction Neutralization Test (PRNT) Accuracy in Evaluating Humoral Immune Response to SARS-CoV-2. Diseases 12 (2024).

39. D. Bates, M. Mächler, B. Bolker, S. Walker, Fitting Linear Mixed-Effects Models Using lme4. J. Stat. Softw. 67, 1–48 (2015).

40. R. C. Team (2022) R: A language and environment for statistical computing. R Foundation for Statistical Computing, Vienna, Austria. https://www.R-project.org/.

